# Preterm and early-term delivery after heat waves in eight US states: A case-crossover study using the High-resolution Urban Meteorology for Impacts Dataset (HUMID)

**DOI:** 10.1101/2025.03.04.25323373

**Authors:** Amy Fitch, Mengjiao Huang, Matthew Strickland, Andrew Newman, Christina Kalb, Joshua L. Warren, Xiaping Zheng, Howard Chang, Lyndsey Darrow

## Abstract

**Background:** Heat wave frequency and intensity is increasing and this trend is more pronounced in urban areas. Heat waves may be acutely associated with early birth.

**Objectives:** To examine the acute relationship between heat waves and preterm (<37 weeks) and early-term (37-38 weeks) birth in eight states: California, Florida, Georgia, Kansas, Nevada, New Jersey, North Carolina, and Oregon

**Methods:** Daily mean temperatures from the novel High-resolution Urban Meteorology for Impacts Dataset (HUMID) were averaged by zip code tabulation area (ZCTA) and linked to singleton preterm and early-term births identified statewide from vital records. Heat waves were defined based on days exceeding the local 97.5%ile temperature threshold during the 4-day exposure window preceding birth. We conducted case-crossover (conditional logistic regression) state-specific analyses and pooled results using inverse-variance weighting to obtain summary effect estimates. We also calculated ORs adjusting for temporal changes in the pregnancy risk set, conducted an analysis excluding medically-induced early-term births, and modeled effects stratified by 97.5^th^ mean temperature threshold categories.

**Results:** The analysis included 2,966,661 early-term and 945,869 preterm births occurring from May - September across the eight states from as early as 1990 to 2017. Results showed modestly elevated odds of early-term birth for heat waves occurring in the 4 days preceding birth. Pooled ORs (95%CIs) for 3- and 4-consecutive days above the 97.5^th^ percentile mean temperature were 1.018 (1.011, 1.026) and 1.017 (1.005, 1.028), respectively. Preterm birth ORs were similar, but less precise; OR=1.015 (1.001, 1.029) and 1.019 (0.999, 1.041) for 3- and 4-consecutive days respectively. Estimated odds ratios tended to be stronger for ZCTAs in the second-lowest category of temperature threshold.

**Discussion:** Using fine-scale surface temperature data capturing urban-heat islands, we observed a modest acute overall effect of heat waves on preterm and early-term birth.

## Introduction

Preterm birth (delivery before 37 weeks’ gestation) occurs in almost 10% of births and contributes to over a third of infant deaths in the United States (US).^1,2^ Preterm birth is also a leading cause of morbidity, including neurological disorders, respiratory disorders, and gastrointestinal disorders,^3,4,5^ and is also associated with cognitive deficits and behavioral disorders.^3,6^ Early-term birth (37-38 weeks’ gestation) is far more common than preterm birth and the proportion of births in these weeks is also increasing, from 24% of births in 2014 to 29% in 2022.^7^ Though health and developmental consequences are generally less severe than preterm, early-term birth is also associated with increased morbidity and mortality during infancy and later in life relative to birth at 39 weeks or later.^8,9,10^

Heat waves are among the most dangerous natural disasters.^11^ Heat is responsible for over 150 deaths per year in the US^12^ and emergency department visits and hospital admissions increased during heat waves.^13^ Cardiovascular, respiratory, and cerebrovascular conditions are the top causes of death associated with heat waves.^14^ A growing body of literature suggests that exposure to extreme heat later in pregnancy increases risk of preterm and early-term birth.^15,16^ In a recent systematic review, four of the five included studies reported a significant association between acute heat exposure and preterm birth^16^ and a 2020 meta-analysis reported a 16% increased risk of preterm birth associated with birth during a heat wave.^15^ While biological mechanisms remain uncertain, extreme heat may overwhelm the body’s ability to thermoregulate, resulting in dehydration and/or inflammation that may ultimately stimulate labor.^17^ Heat waves are increasing in frequency, intensity, and duration, and the heat wave season is lengthening so associated health risks will likely worsen without mitigating interventions.^18^

Studies analyze a variety of heat metrics, some model the effect of continuous temperature^19,20^ and others use categorical measures (e.g., ≥2 days above a threshold temperature).^21,22^ In addition, heat may be measured as minimum, mean, and/or maximum temperature^21,22^ or a related measure such as apparent temperature, accounting for humidity.^20,21,23,24^ Further, most previous studies were conducted with birth data from a single city or state.^19,21,23,24^ Fewer studies address the outcome of early-term birth, with some reporting modest 1-4% relative increases in risk with exposure to heat waves^22^ and others reporting stronger effects, up to a 27% increase.^20,25^

Our study estimates the effect of heat wave exposure in the four days preceding birth (lag 0-3) on preterm (28-36 weeks) and early-term (37-38 weeks) birth. Using a time-stratified case-crossover design, we answer the question: ‘is there an acute effect of heat wave days on early birth relative to other days typical in the same month?’ We use a novel high-resolution meteorological dataset and birth data from eight US states, covering approximately 30% of the US population, with 14-28 years of birth records per state.

## Methods

### Study population

Data for live, singleton births were collected from state vital records in eight US states: California, Florida, Georgia, Kansas, Nevada, New Jersey, North Carolina, and Oregon. These states and years were chosen in order to achieve a large sample size with some geographic diversity and because obtaining birth data was logistically and financially feasible. Our study population includes infants born preterm (28-36 weeks) or early-term (37-38 weeks). We excluded extremely preterm births (20-27 weeks; ∼6% of preterm births) because of their stronger association with intrauterine infection and congenital anomalies and to be consistent with our previous work.^22,26–28^ We excluded births if the maternal state of residence did not match the birth state or the maternal zip code was missing or did not match a 2010 US Census Zip Code Tabulation Area (ZCTA) in the state (approximately 1.5% of births). The study population included births from 1990 through 2017, or a subset of these years depending on state (see Table 1).

**Table 1:**
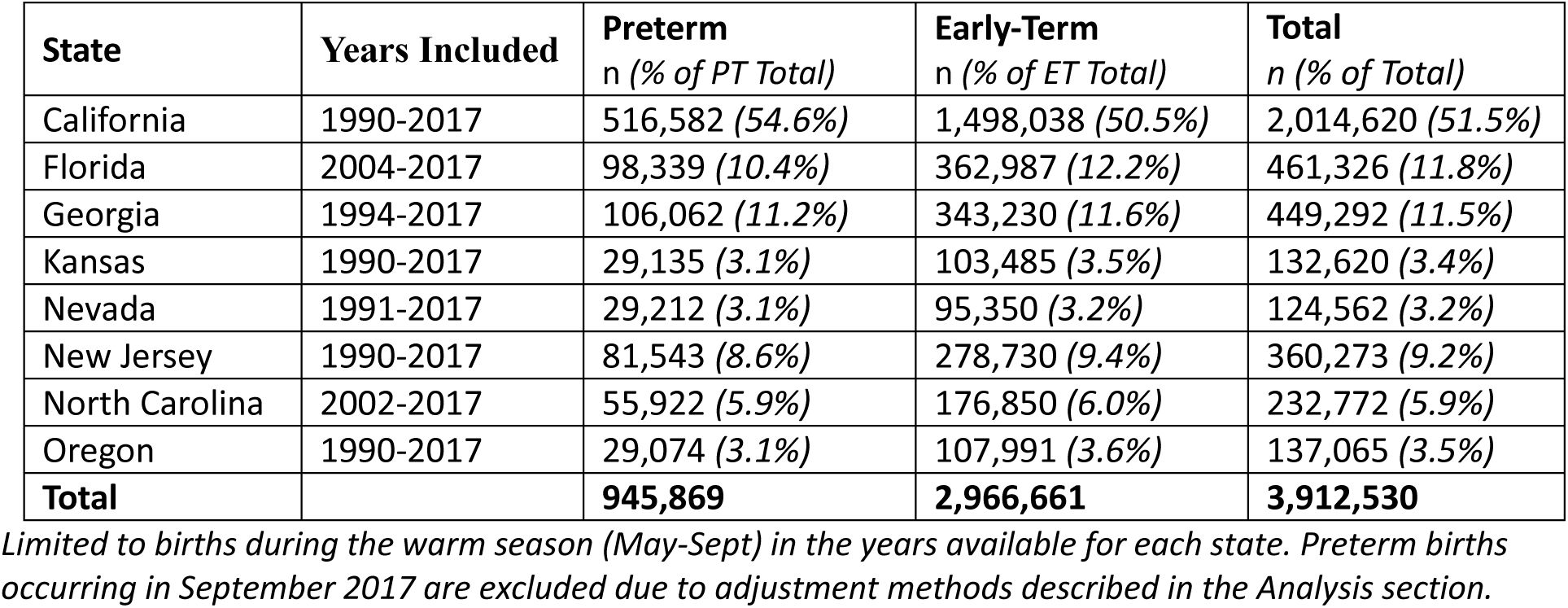
Number and percent of preterm and early-term births included in the analyses for each state.

### Study design

We used a time-stratified, case-crossover design, which inherently controls for all time-invariant factors as participants serve as their own controls.^29^ Case-crossover is a case-only design where participants serve as their own controls, inherently accounting for time-invariant confounders. Numerous studies have established the appropriateness of this study design for this area of research when accounting for any within-month trends.^30–33^ Presence or absence of a heat wave was determined for exposure periods linked to the case day (day of birth) and three or four control days in the same calendar month, matched on day of the week. The 4-day exposure periods were defined by the day of delivery (or control day, lag 0) and the three previous days (lag1, lag2, and lag3). We selected the 4-day exposure window *a priori* based on our previous study showing stronger effects for a more acute 4-day window compared to a 7-day window.^26^ The analysis was limited to births occurring during the warm season, May 1 through September 30. For the preterm outcome, September 2017 was excluded to allow for full enumeration of the risk set necessary for our analysis methods, described in more detail in the Analysis section below.

### Environmental exposure

We obtained 1 km x 1 km gridded estimates of daily maximum and minimum temperatures from the novel High-resolution Urban Meteorology for Impacts Dataset (HUMID).^34,35^ We used this novel dataset to better account for spatial variability in temperatures due to urban heat islands. HUMID was designed to provide an improved spatial representation of temperature distribution across urban areas than existing meteorological datasets by incorporating an explicit urban canopy model into a land-surface modeling system. This system uses meteorological inputs such as solar radiation, air temperature, precipitation, wind, and static geophysical attributes such as soil type, vegetation, as well as the percent urban fraction from the National Land Cover Database^36^ to estimate the full energy and water balances at the surface over natural and built environments. Further, a bias correction process was applied to air temperature using available station observations to correct model errors yet was designed to prevent over-smoothing of high-frequency spatial variability in the model-observation fusion.^34^ HUMID appears to better resolve spatial temperature variability within urban areas and across urban-rural divides as compared to other gridded meteorology products which do not explicitly account for urban areas and may only implicitly resolve portions of the urban areas due to interpolation techniques and input station densities^34^ and supports our spatially detailed exposure estimates at the ZCTA level. For each day of the study period we averaged daily mean temperatures among grid points that fall within ZCTA polygon boundaries, based on 2010 US Census ZCTA state shapefiles, in order to assign exposure based on the maternal residence information in the birth records.^37^

#### Heat wave definitions

We defined heat waves using daily mean temperature and a 97.5^th^ percentile threshold calculated at the ZCTA-level based on the full 28-year study period. We used a location-specific relative threshold to account for acclimatization^38^ and chose the 97.5^th^ percentile in order to explore the effect of more extreme events. As there is no universally accepted definition of a heat wave, we used several definitions to capture different ways of looking at heat waves. We selected the three heat wave definitions *a priori* to be consistent with our previous research.^22,26^

- Heat wave definition 1 (HW1): The number of days over the 97.5^th^ percentile threshold in a four-day period, whether or not the hot days are consecutive. (1 to 4 days; reference = 0)
- Heat wave definition 2 (HW2): The number of *consecutive* days over the 97.5^th^ percentile threshold in a four-day period (≥2-, ≥3- or 4-consecutive days; reference = <2-, <3-, or <4-consecutive days, respectively). This definition emphasizes heat wave duration.
- Heat wave definition 3 (HW3): The average degrees exceeding the 97.5% percentile in a four-day period, calculated as the four-day average temperature minus the 97.5^th^ percentile temperature (included as a linear term). If negative, the value is set to zero. Two exposure windows meeting the same metrics for definitions 1 and 2 would differ on their HW3 value if the hot days in one window are hotter than the days in the other window. This definition emphasizes heat wave intensity.

### Analysis

We estimated state-specific odds ratios using conditional logistic regression (SAS 9.4). State-specific results were pooled using inverse-variance weighting (fixed-effects meta-analysis) in order to present a summary measure of the common effect in the eight states (R 4.3.2). Under certain conditions, the odds ratios from these models could be interpreted as rate ratios.^39^ All births were included in the primary analyses due to uncertainty about biological mechanisms, but we conducted a sensitivity analysis for the early-term outcome where medically-induced births were excluded for the four states where induction data were available: California, Kansas, Nevada, and Oregon. We also explored whether associations were modified by differences in climate within and across states. To do this we separated ZCTAs into four categories based on the 97.5^th^ percentile mean temperature threshold: T1: <27°C (<80.6°F), T2: 27 – 29°C (80.6 – 85°F), T3: 30 – 31°C (86 – 89.5°F), and T4: ≥ 32°C (≥ 89.6°F). Category cut points were chosen before analysis to yield similarly sized groups.

In addition to the standard case-crossover analysis, we also calculated adjusted odds ratios by adapting an approach developed for time-series analysis by Vicedo-Cabrera et al.^24^ The adjustment variable (*Wi*) is the average probability of birth on each day in each ZCTA among all ongoing gestations at risk of the outcome (28-36 weeks for preterm and 37-38 weeks for early-term). The goal of the adjustment is to minimize bias resulting from within-window changes in risk of the outcomes due to seasonal trends in conception^33,40^ as well as within-window trends in misclassification of gestational age due to preferential reporting of the last menstrual period as the 15^th^ of the month.^41,42^ The adjustment term *Wi* for preterm birth in week *i* was calculated as 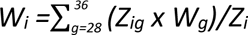 where *Zig* is the ZCTA-level count of fetuses at risk of preterm birth at gestational week *g* during calendar week *i*; *Zi* is the ZCTA-level count of all fetuses during calendar week *i*; and *Wg* is the probability of birth at each gestational week *g*, calculated from each state’s complete birth records for the study period. The adjustment formula for early-term birth is similar, with different considered gestational weeks: 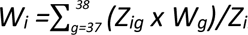 Validity of this approach in this case crossover context is supported by previous simulation and discussed in more detail in the supplement.

For the preterm birth analysis, all gestations at risk of the outcome could not be enumerated after September 3, 2017 in the absence of 2018 birth data. For this reason, the preterm analysis excludes September 2017. This was not an issue for the early-term analysis because the risk period is shorter and the unenumerated at-risk gestations fell outside the warm season.

## Results

### Descriptive data

A total of 945,869 preterm births and 2,966,661 early-term births across the eight states were included in the analysis. This includes births during the warm season (May – September) for the years available, where maternal residence in the birth record could be linked to a ZCTA in the US Census 2010 state shapefile.^37^ Across the eight states, an average of 98% of preterm and early-term births met these criteria. Maternal characteristics of the study population for preterm and early-term births are shown in Table 2. Maternal characteristics broken down by state are available in Table S1.

**Table 2:** Maternal Demographics of cases.

| Total Cases | Preterm<br>945,869 |  | Early Term<br>2,966,661 |  |
| --- | --- | --- | --- | --- |
| Age | N | % | N | % |
| 10-24 | 332,171 | 35.12 | 927,663 | 31.3 |
| 25-34 | 453,049 | 47.90 | 1,542,178 | 52.0 |
| 35-55 | 160,502 | 16.97 | 496,611 | 16.7 |
| Unknown/Implausible | 147 | 0.02 | 209 | 0.0 |
| Race/Ethnicity |  |  |  |  |
| White non-Hispanic | 336,678 | 35.59 | 1,170,495 | 39.5 |
| Black non-Hispanic | 160,868 | 17.01 | 391,151 | 13.2 |
| Hispanic | 348,990 | 36.90 | 1,067,943 | 36.0 |
| Other | 91,898 | 9.72 | 314,802 | 10.6 |
| Unknown | 7,435 | 0.79 | 22,270 | 0.8 |
| Education* |  |  |  |  |
| Less than high school | 246,179 | 27.66 | 645,313 | 23.1 |
| High school/GED | 267,605 | 30.07 | 799,850 | 28.7 |
| More than high school | 357,362 | 40.16 | 1,292,956 | 46.3 |
| Unknown | 18,801 | 2.11 | 51,692 | 1.9 |

*\*These categories of educational achievement data not available in North Carolina birth records (preterm n = 889,947; early-term n = 2,789,811).*

Table 3 shows the proportion of total case days and control days in each birth analysis (preterm and early-term) that met each heat wave definition. The heat wave event days made up between 1% and 7% of the case and control days in the analysis, depending on the heat wave metric and birth outcome. The state-specific proportion of case days and control days that met each heat wave definition is available in Table S2.

**Table 3:** Percent of case days and control days meeting each heat wave definition using a four-day (lag 0-3) exposure window.

|  | **Preterm** |  | **Early Term** |  |
| --- | --- | --- | --- | --- |
|  | Case Days | Control Days | Case Days | Control Days |
| Total Days | 945,869 | 3,244,287 | 2,966,661 | 10,156,872 |
| **HW1** |  |  |  |  |
| 1 day | 5.54 | 5.50 | 5.68 | 5.67 |
| 2 days | 3.48 | 3.48 | 3.55 | 3.56 |
| 3 days | 1.98 | 1.99 | 2.04 | 2.04 |
| 4 days | 1.34 | 1.33 | 1.37 | 1.37 |
| **HW2** |  |  |  |  |
| ≥ 2 days | 6.50 | 6.51 | 6.66 | 6.67 |
| ≥ 3 days | 3.08 | 3.08 | 3.16 | 3.16 |
| 4 days | 1.34 | 1.33 | 1.37 | 1.37 |
| **HW3** |  |  |  |  |
| % days > 0 | 4.25 | 4.24 | 4.38 | 4.38 |
| Mean °C* | 0.91 | 0.92 | 0.90 | 0.91 |

\*Mean degrees Celsius above the 97.5<sup>th</sup> percentile among days where HW3>0

### Outcomes

#### Preterm

Odds ratios (OR) and 95% confidence intervals (CI) for heat waves and preterm birth are shown in Figure 1 with pooled estimates at the top and state-specific results below. Pooled odds ratios showed modest positive associations for heat waves occurring in the four days preceding birth. The estimates tended to be stronger with more days and more consecutive days of heat wave exposure and there was an increase in the odds of preterm birth per degree in the average temperature above the 97.5^th^ percentile threshold.

**Figure 1:**
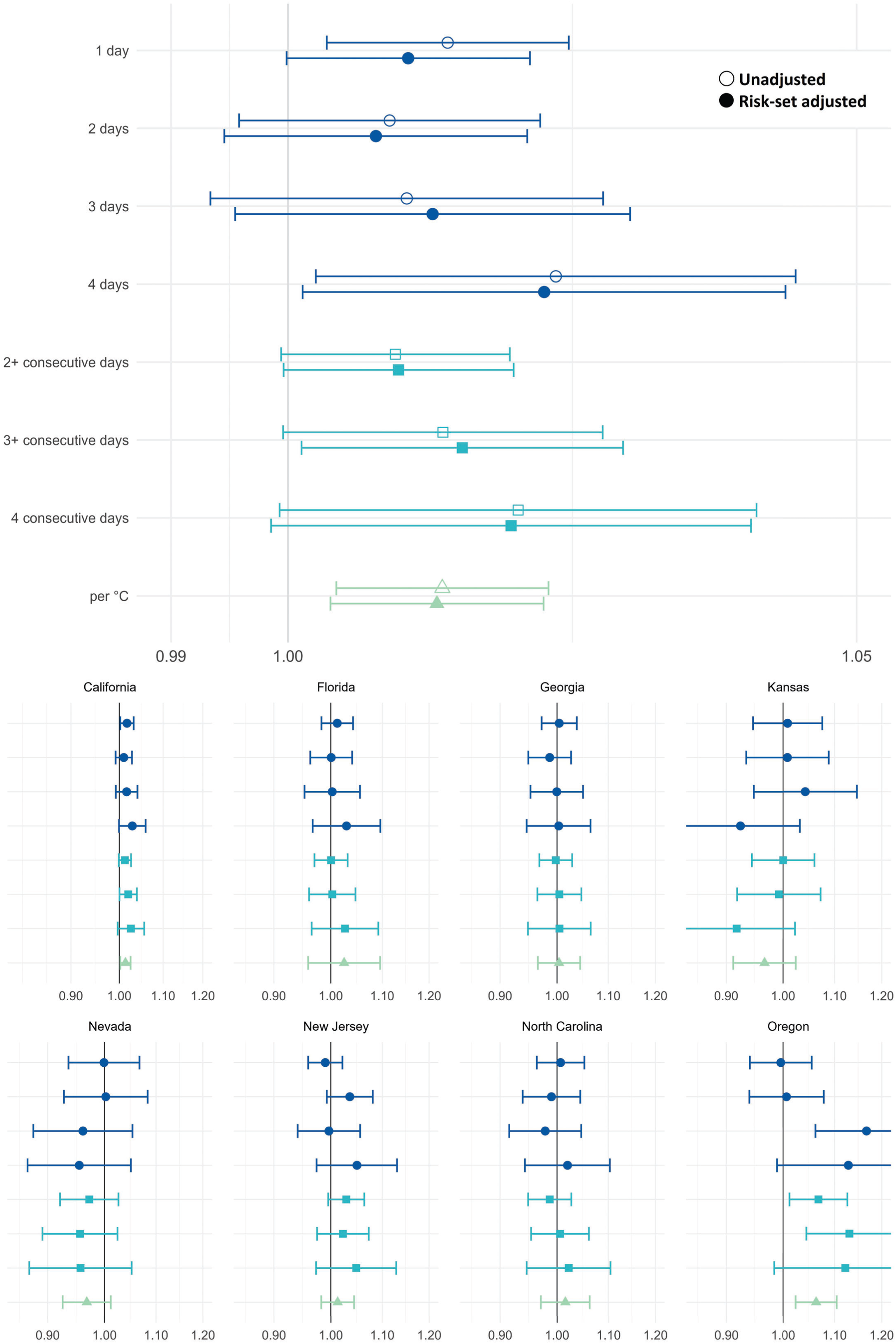
Pooled odds ratios (top panel) and odds ratios by state (bottom panel) and 95% confidence intervals for the association between heatwaves and preterm birth in California, Florida, Georgia, Kansas, Nevada, New Jersey, North Carolina, and Oregon. Note the scale difference in state-specific and pooled results. Standard case-crossover analysis results are shown in outlined shapes and results adjusted for probability of preterm birth among gestations at-risk are shown in solid shapes. The reference category for heatwave definition 1 (dark blue circles) is 0 hot days in the previous week. Heatwave definition 2 (blue squares) are dichotomous exposure categories where the reference categories are <2-, <3-, and <4-consecutive days, respectively. Heatwave definition 3 (green triangle) represents the odds ratio associated with a 1°C increase in the 4-day average degrees over the 97.5th percentile. Estimates are shown in Table S3.

The adjustment for average probability of preterm birth among the risk set in the ZCTA did not meaningfully affect the estimates; for example, the pooled adjusted odds ratio for a <u>≥</u>3-consecutive day heat wave was 1.015 (95% confidence interval: 1.001, 1.029) vs. 1.013 (95% CI: 1.000, 1.027) for the standard (unadjusted) model. Nonetheless, we focus on reporting the adjusted results below because they account for the changing risk of early birth over the month, resulting from seasonal trends in conception and measurement error due to preferential reporting of last menstrual period dates.

The adjusted pooled odds ratios across heat wave metrics ranged from 1.008 to 1.022 for preterm birth. For every 1°C increase in the average temperature above the threshold over the four-day exposure window, the odds of preterm birth increased by 1.3% (95% CI: 1.004, 1.022). The largest increase in preterm birth risk was seen when all four days were above the threshold compared to zero days, OR=1.022 (95% CI: 1.001, 1.044). Full numerical results are presented in Table S3.

Adjusted state-specific preterm results (bottom of Figure 1) show some variability across the eight states, although confidence intervals overlapped across states and most results were not statistically significant (p < 0.05). California’s results are the most precise suggesting small increases in preterm birth following heat waves, with ORs ranging from 1.010 to 1.029. In a post-hoc sensitivity analysis excluding California, pooled results for the remaining seven states followed a similar pattern, but were less precise due to smaller sample size (see Figure S1). The magnitude of associations was notably highest in Oregon where exposure to three hot days in the four days before birth (vs. zero) is associated with a 16.7% increase in the odds of preterm birth (95% CI: 1.062, 1.282). However, some states, for example Nevada, had more negative point estimates than positive. All state-specific estimates for both outcomes and for both standard and adjusted models are presented in Table S3.

#### Early-Term

Similar to preterm birth, pooled adjusted odds ratios for early-term birth showed modest positive associations for heat waves occurring in the four days preceding birth. Pooled odds ratios ranged from 1.007 to 1.020 across the heat wave metrics. Results for the pooled early-term analyses were more precise than the preterm models due to higher outcome counts. The strongest associations were seen for the 3- and 4-day heat wave compared to zero days, OR=1.020 (95% CI: 1.010, 1.030) and OR=1.020 (95% CI: 1.008, 1.032), respectively. For every 1°C increase in average temperature above the 97.5^th^ percentile threshold the odds of early-term birth increased by 1.0% (OR=1.010, 95% CI: 1.005, 1.016).

Adjusted state-specific results for early-term birth are shown in the lower portion of Figure 2 and, like preterm birth results, estimates vary across states. The most consistent positive and precise associations were seen in California, with ORs ranging from 1.017 to 1.035, all but one of which are statistically significant. Negative associations are seen in Kansas and North Carolina, but the results are relatively imprecise and almost all are non-significant. The post-hoc sensitivity analysis excluding California yielded less consistent evidence for a positive association (see Figure S1).

**Figure 2:**
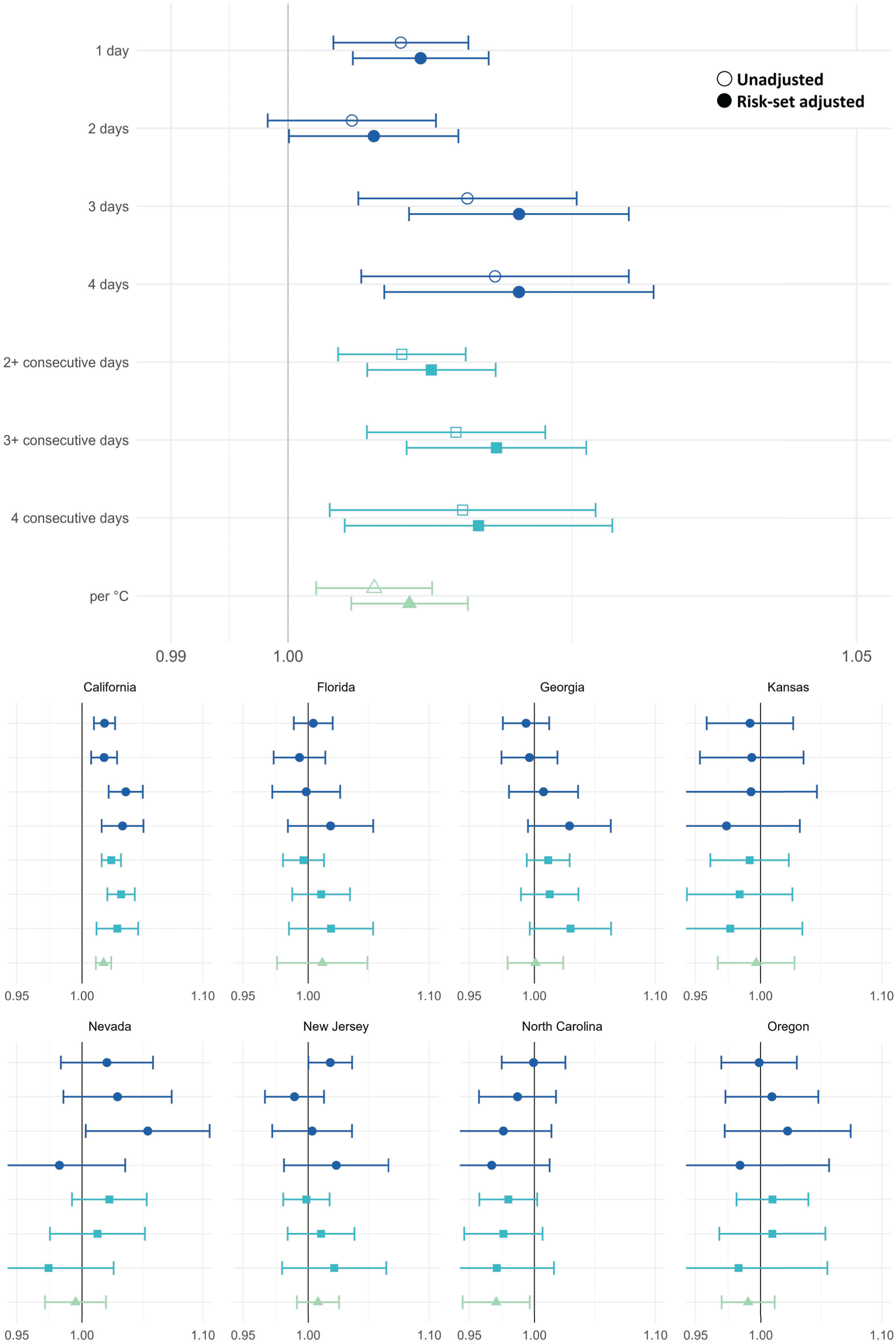
Pooled odds ratios (top panel) and odds ratios by state (bottom panel) and 95% confidence intervals for the association between heatwaves and early-term birth in California, Florida, Georgia, Kansas, Nevada, New Jersey, North Carolina, and Oregon. Note the scale difference in state-specific and pooled results. Standard case-crossover analysis results are shown in outlined shapes and results adjusted for probability of preterm birth among gestations at-risk are shown in solid shapes. The reference category for heatwave definition 1 (dark blue circles) is 0 hot days in the previous week. Heatwave definition 2 (blue squares) are dichotomous exposure categories where the reference categories are <2-, <3-, and <4-consecutive days, respectively. Heatwave definition 3 (green triangle) represents the odds ratio associated with a 1°C increase in the 4-day average degrees over the 97.5th percentile. Estimates are shown in Table S3.

#### Sensitivity analysis

For the four-state analysis of early-term births excluding medically-induced births (11% excluded), the odds ratios are similar to the primary early-term analysis, see Figure S2. For example, for every 1°C increase in 4-day average temperature above the threshold the pooled, adjusted OR excluding inductions was 1.015 (95% CI: 1.009, 1.021) vs. 1.013 (95% CI: 1.008, 1.019) for the full four-state population.

#### Stratification by temperature threshold

To determine if the effect varies by differences in climate, we conducted analyses stratified by 97.5^th^ percentile mean temperature threshold level applied to each ZCTA. Results of the stratified analysis are shown in Figure 3. There were minimal differences among the groups and confidence intervals generally overlap across categories; however, the T2 group (thresholds from 27-29°C) has the most consistently elevated odds ratios for both preterm early-term birth. The T4 group representing the hottest locations in our analysis (including Las Vegas, southern inland California, and Miami, see Figure S3) showed the least evidence for an association for the most extreme heat wave definitions.

**Figure 3:**
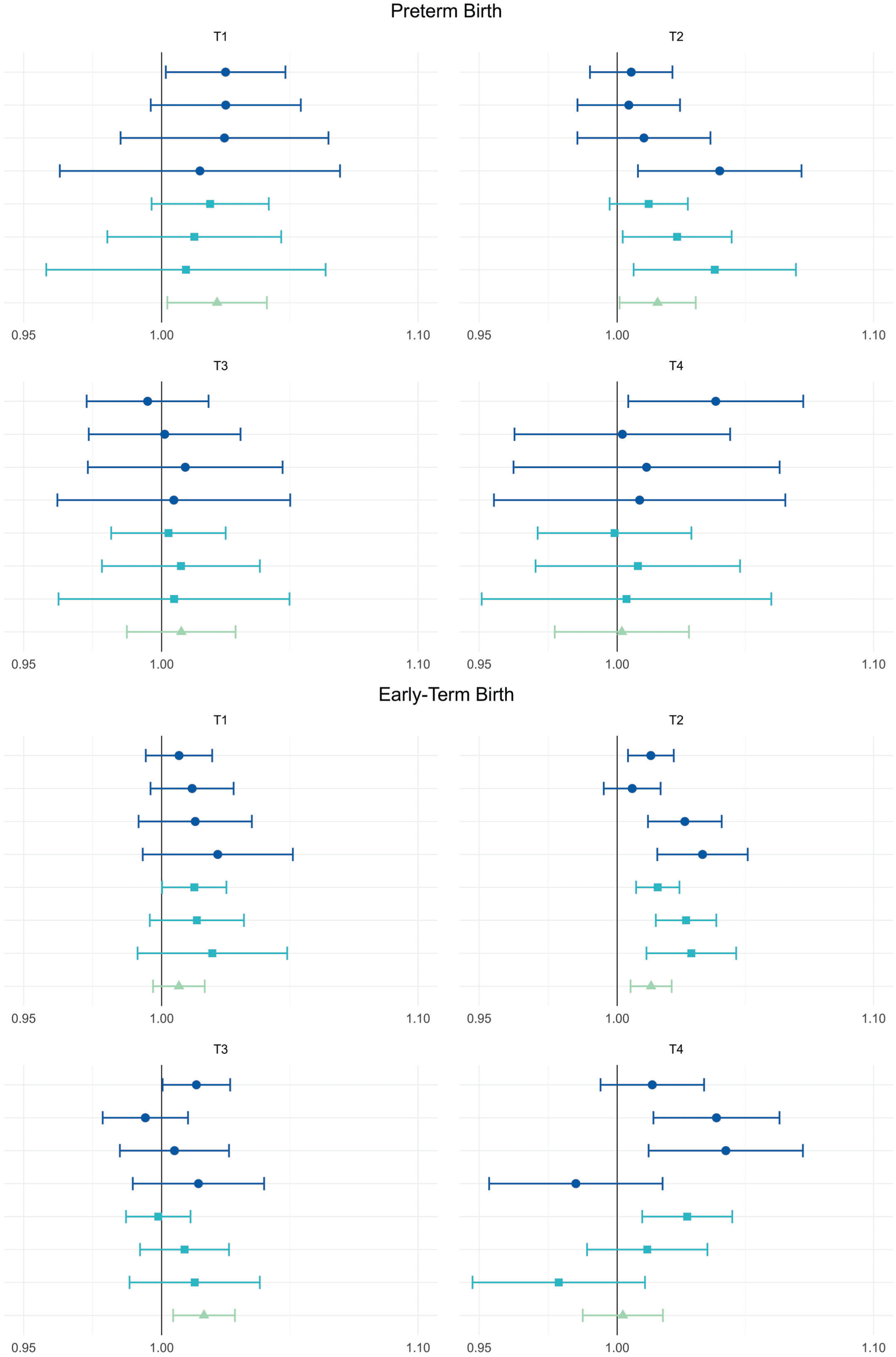
Pooled odds ratios and 95% confidence intervals for the association between heatwaves and preterm birth (top panel) and early-term birth (bottom panel), stratified by 97.5th percentile mean temperature range. T1: <27°C (<80.6°F) includes CA, GA, NV, NJ, NC, & OR; T2: 27 – 29°C (80.6 – 85°F) includes all eight states; T3: 30 – 31°C (86 – 89.5°F) includes CA, FL, GA, KS, NV, NJ, & NC; and T4: ≥ 32°C (≥ 89.6°F) includes CA, FL, GA, KS, NV, and NC. The reference category for heatwave definition 1 (dark blue circles) is 0 hot days in the previous week. Heatwave definition 2 (blue squares) are dichotomous exposure categories where the reference categories are <2-, <3-, and <4-consecutive days, respectively. Heatwave definition 3 (green triangle) represents the odds ratio associated with a 1°C increase in the 4-day average degrees over the 97.5th percentile. Estimates are shown in Table S4.

## Discussion

In this study of births in eight states from 1990 to 2017, we found a small elevated risk of preterm and early-term birth acutely following heat wave events. We applied a consistent approach to a large study population (including 945,869 preterm births and 2,966,661 early-term births), using high spatial-resolution temperature models to assign temperatures at the ZCTA level, and we pooled state-specific results into summary odds ratios to precisely estimate the effect of a historically rare exposure. We were motivated to examine multiple heat wave definitions, in part due to the lack of a gold standard definition, and also to explore if different dimensions of heat waves (duration and intensity) affect associations. Taking all the metrics examined together, there was a trend of stronger associations for longer duration and higher intensity heat waves, and results were generally consistent across the different definitions. These results can be interpreted as the effect of extreme ambient temperatures as an acute trigger for early birth, after any adaptive behaviors have occurred (e.g., reducing outdoor activity, utilization of air conditioning).

The positive associations for both preterm and early-term outcomes were heavily influenced by California, which accounted for 55% of preterm and 50% of early-term births in the respective analyses. A sensitivity analysis excluding California showed that California was particularly influential on the early-term birth results, whereas the preterm results excluding California show similar overall pattern of positive associations with less precision.

Most previous studies using time-series or case-crossover methodology report acute increases in preterm birth risk ranging from 2 - 20%.^19–21,23,24^ Varied effect sizes have been observed within studies depending on the heat wave definition. For example, hazard ratios in one California study range from 1.008 for a two-day heat wave using a 75^th^ percentile threshold to 1.128 for four-day heat waves at the 98^th^ percentile threshold.^21^ In general, the magnitude of observed associations in our study was smaller than most previous studies, which could be explained by differences in study population, design, or exposure definitions (or a combination). In previous studies, exposure has been defined based on continuous temperature rather than heat wave metrics,^19,43^ apparent temperature rather than temperature,^43^ and different percentile thresholds, often less extreme than the 97.5^th^ percentile applied in our study.^44^ Our early-term results are most similar to those of Ilango et al.,^14^ covering similar years in a California population using a survival analysis approach, which also accounts for the gestational age of the risk set. This alignment makes sense since California was highly influential in our analysis. Our preterm results, however, are weaker relative to the same study. However, the magnitude of our pooled results is consistent with those estimated in studies of the 50 most populous US cities using the same heat wave definitions but less spatially refined temperature estimates.^22,26^ Prior work showing similar results for temperature and apparent-temperature based metrics suggests this is unlikely to explain differences between studies, at least for studies leveraging within-location, acute, temporal exposure contrasts such as this one.^22,45^

Some studies compare individuals while our comparison is within-person, removing any potential confounding by individual characteristics. The monthly time-stratified design also inherently controls for long-term meteorological trends (increasing heat wave frequency) and trends in birth outcomes (declining preterm birth rates) because the comparisons occur within a single month. Our design resulted in very tight control for seasonal and long-term trends, but this was at the cost of exposure variability as temperature changes within a calendar month are less pronounced than across months and across space. Our comparison of heat wave days to other days typical of that month may attenuate associations compared to other studies with different referents. Approaches to selection of control days in case-crossover studies can vary^20^ and some have been shown to introduce considerable positive bias.^33^ In addition, previous case-crossover analyses lack adjustment for the expected count of early births based on count of pregnancies at risk and probability of birth among them. While the adjustment did not meaningfully change our results, it may be more necessary to obtain unbiased estimates in studies that assess continuous temperature, which shows more systematic within-month trends than heat waves as defined in our study.

Estimates in our study were not consistent across states or even within states across heat wave metrics, though we note that most results were imprecise as expected based on the relatively infrequency of heat waves, occurring on only 1.3-6.5% of case or control days. In California, which contains half of the study population, there was a positive trend with stronger associations seen for longer duration heat waves. California’s HW3 results showed a 1.4% and 1.7% increased risk of preterm and early-term birth, respectively, for every 1°C increase in 4-day average temperature above the threshold. As expected, smaller states show more variable and less precise estimates ranging from strong positive associations (preterm results in Oregon) to estimates below 1.0 (preterm birth results in Nevada and early-term birth results in North Carolina and Kansas), but confidence intervals largely overlap. We designed the study to calculate precise overall pooled estimates results, and with only 8 states our ability to distinguish heterogeneity from random error was limited. Nonetheless, true variation in the effect of heat waves on early term birth may be expected based on population-specific characteristics and behaviors and could be explored in future multi-site studies.

It is likely that the acute effect of heat waves on early birth is modified by protective behaviors during the most extreme heat events and/or differences in adaptive capacity (e.g., availability or utilization of air conditioning). Air conditioning prevalence is associated with reduced heat-related mortality^46^ and may also modify the association between heat and birth outcomes. In 2020, 91% of North Carolina households and 92% of Nevada households reported using air conditioning, well above California’s 72% and Oregon’s 76%,^47^ which may explain why our estimates for California and Oregon were more in the positive direction (air conditioning prevalence was also high in the other four states, 93-96%). In our study, where exposure contrasts are made within a calendar month, we are often comparing extremely hot days to referent very hot days. In this situation, differences in adaptive capacity and protective behaviors could result in an apparent protective effect rather than a null effect if the referent days in the same month were hot enough to cause an increased risk of early birth, but they were not hot enough to prompt behavior changes that occur during the longest and hottest heat wave days. Liang et al. report a protective effect in their study based in Shenzhen, China, which they attribute to high prevalence of air conditioning in the high socio-economic status city.^48^

Several of the previous studies that find the largest effects were conducted in Europe where heat waves have historically caused much higher mortality than the US. Since 1979, a total of approximately 11,000 Americans have died from heat related illnesses, including 1,250 in a 1995 heat wave, considered the most significant heat wave event in modern US history.^49^ Contrast that with Europe where more than 60,000 deaths in a single year (2022) were attributed to extreme heat.^50^ Although air conditioning use has increased in Europe in recent years, it is still far less common throughout Europe than it is in the US. Globally, the US is second only to China in sales of residential air conditioning units and household ownership of air conditioning is over 90% in the US, compared to less than 10% in Europe. Such geographic differences in air conditioning utilization and perhaps other aspects of adaptive capacity should result in differences in maternal experienced heat, even when ambient air temperatures and heat wave metrics are similar. For these reasons, US populations are expected to exhibit weaker overall population-level associations between heat waves and health outcomes than other areas.

We found that in some geographic areas, such as the California coast, there was little variability in the mean temperature throughout the year. In these places the 97.5^th^ percentile threshold temperatures were relatively low, leading to the possibility that days classified as heat wave days are not capturing the exposure of interest. The mechanism to explain an acute association between heat wave exposure and pre-/early-term birth is not fully understood, but exogenous heat may trigger early labor as a result of reduced uterine blood flow caused by dehydration and/or inflammation associated with maternal production of heat shock proteins. In an area with a comfortably low 97.5^th^ percentile temperature threshold, temperatures exceeding that threshold may not be hot enough to trigger the biological mechanism. This concern motivated the sub-analysis stratifying by category of absolute temperature threshold. Although results were imprecise, the associations between heat waves and the birth outcomes tended to be highest for the middle (T2) temperature group compared to the hottest and the coolest ZCTAs. The hottest ZCTAs (T4, including Las Vegas, southern inland California, and Miami, as shown in Figure S3) showed no evidence of an elevated association for the longest duration and most intense heat waves, possibly reflecting increased use of air conditioning and/or behavior change during the most extreme events.

### Limitations

It is possible that some of the geographic variation in our results could be due to differing levels of air pollution. Air pollution may mediate the association between heat waves and early birth, as extreme heat can exacerbate poor air quality and certain air pollutants are associated with preterm birth risk. Additionally, in particular in the western states, wildfires that co-occur with heat waves could play a role in these associations.^54^

While, the HUMID dataset provides high-resolution temperature estimates, we calculated averages at the ZCTA level to align with residence information provided in the birth records. Such aggregation reduces the precision of the exposure measurement, but may also better capture maternal heat exposure as pregnant people spend the majority of the day “at or near” home later in pregnancy.^55^

We acknowledge there is some measurement error of gestational age in the birth records and that the magnitude of these errors has changed over time as clinical methods have evolved. These concerns are mitigated by our focus on acute effects, where we use within-month exposure contrasts. We don’t believe the outcome errors would result in spurious associations as they are independent of short-term variations in the exposure.

Finally, while our study has a large sample size, we have data for only eight states and our results may not be generalizable to all states or countries. Results were somewhat variable across our states, indicating that the presence or magnitude of the risk may vary depending on regional climates or other factors with spatial variability. However, our results are consistent with our previous work in the 50 most populous cities across the US.^22,26^

### Conclusion & Future Directions

Our study evaluates overall average population-level effects, possibly obscuring more vulnerable subgroups who may experience greater effects. Unfortunately, spatially resolved air conditioning data are not available in the US, preventing us from effectively exploring that potential effect modifier. Despite mitigating factors (protective behaviors, relatively high access to air conditioning, etc.), our eight-state study of almost one million preterm and almost three million early-term births utilizing fine scale temperature data suggests a modest overall population-level acute association between heat waves and early delivery.

Future research in this area should explore vulnerabilities, in particular access to and utilization of AC. These data are not available at a fine spatial scale across multiple states; such data sources need to be developed. In addition, future work should explore maternal behavior changes during heat waves, how those adaptations may mitigate perinatal risks, and how to encourage or support such adaptation.

## Data Availability

Meteorology data are available at:
https://rda.ucar.edu/datasets/d314008/
Birth record data must be obtained directly from state health departments

## Notes

The authors declare they have no conflicts of interest related to this work to disclose.

### Competing Interest Statement

The authors have declared no competing interest.

### Funding Statement

The study was funded by The National Institute of Environmental Health Sciences (NIH) grant # R01ES028346

### Author Declarations

The University of Nevada, Reno Institutional Review Board gave ethical approval for this work.

