## Supplemental Material 1 for "Preterm and early-term delivery after heat waves in eight US states: A case-crossover study using the High-resolution Urban Meteorology for Impacts Dataset (HUMID)"

**Section A**: Adjustment for average probability of birth among gestations at risk

**Figure S1:** Pooled odds ratios and 95% confidence intervals for the association between heatwaves and early-term birth, including and excluding California.

**Figure S2:** Pooled odds ratios and 95% confidence intervals for the association between heatwaves and early-term birth, including and excluding medically-induced births in California, Kansas, Nevada, and Oregon.

**Figure S3:** Geographic distribution of 97.5$^{th}$ percentile temperature categories (T1-T4) by ZCTA

**Section A:** Adjustment for average probability of birth among gestations at risk:

The adjustment term combines the number of ongoing pregnancies at each gestational age and the probability of birth at that gestational age (calculated based on the full state population) to obtain the average probability of birth among ongoing pregnancies at risk. We recently demonstrated via simulation that the risk-set adjustment approach does correct a small positive bias in case-crossover analyses of temperature and preterm birth.[23] Vicedo-Cabrera et al. and Huang et al. both included *log(*$W_i$*) in their models*; however, we chose to include the unlogged *Wi* value because our dataset included many ZCTAs/dates with zero pregnancies at risk. $W_i$ represents the average probability of preterm or early-term birth among ongoing gestations in that ZCTA on that day. We repeated the warm season simulation analysis conducted in Huang et al. adjusting for $W_i$ instead of *log(*$W_i$*)* and found that this also eliminated the small positive bias. See the box plot on the right showing unbiased estimation of a simulated null effect of temperature after adjusting for Wi; the unadjusted estimate on the left shows the expected positive bias using an unadjusted time-stratified case-crossover approach (as shown in Huang et al. 2023).

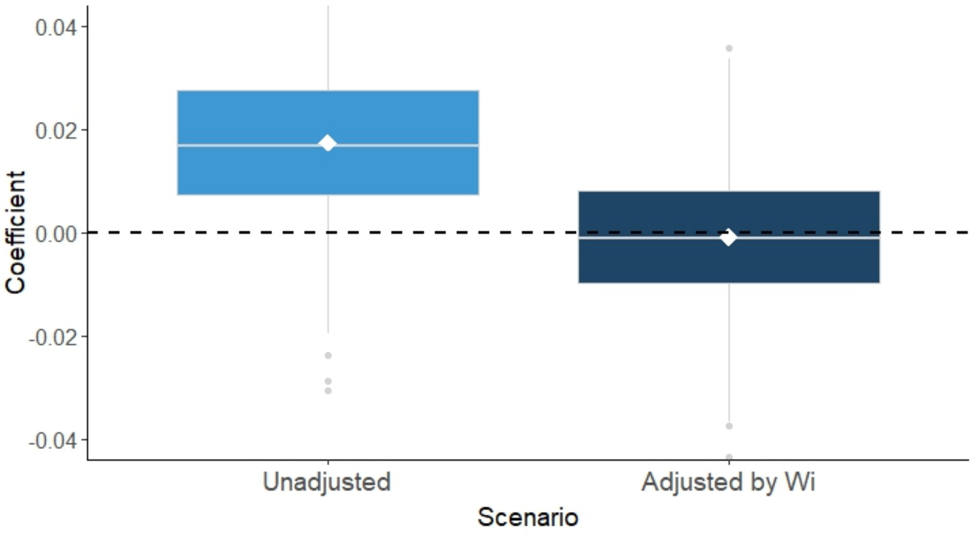


DAG showing confounding by gestational age of the risk set.

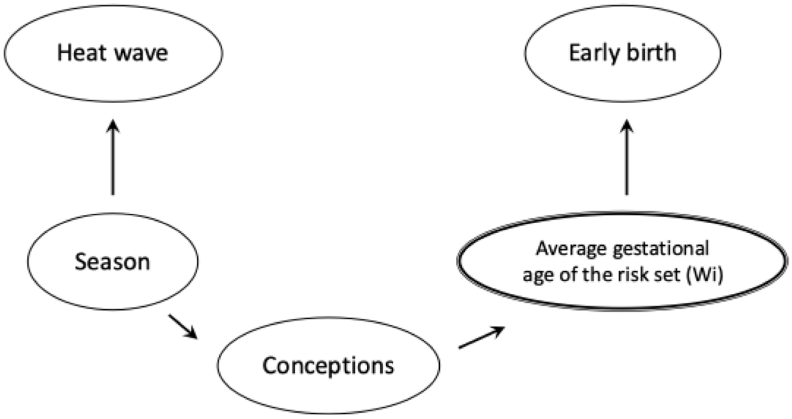


Vicedo-Cabrera AM, Iñíguez C, Barona C, Ballester F. Exposure to elevated temperatures and risk of preterm birth in Valencia, Spain. *Environ Res*. 2014; 134:210-217. doi:10.1016/j.envres.2014.07.021

Huang M, Strickland MJ, Richards M, Warren JL, Chang HH, Darrow LA. Confounding by conception seasonality in studies of temperature and preterm birth: A simulation study. *Epidemiology*. 2023;34(3):439-449. doi:10.1097/EDE.0000000000001588

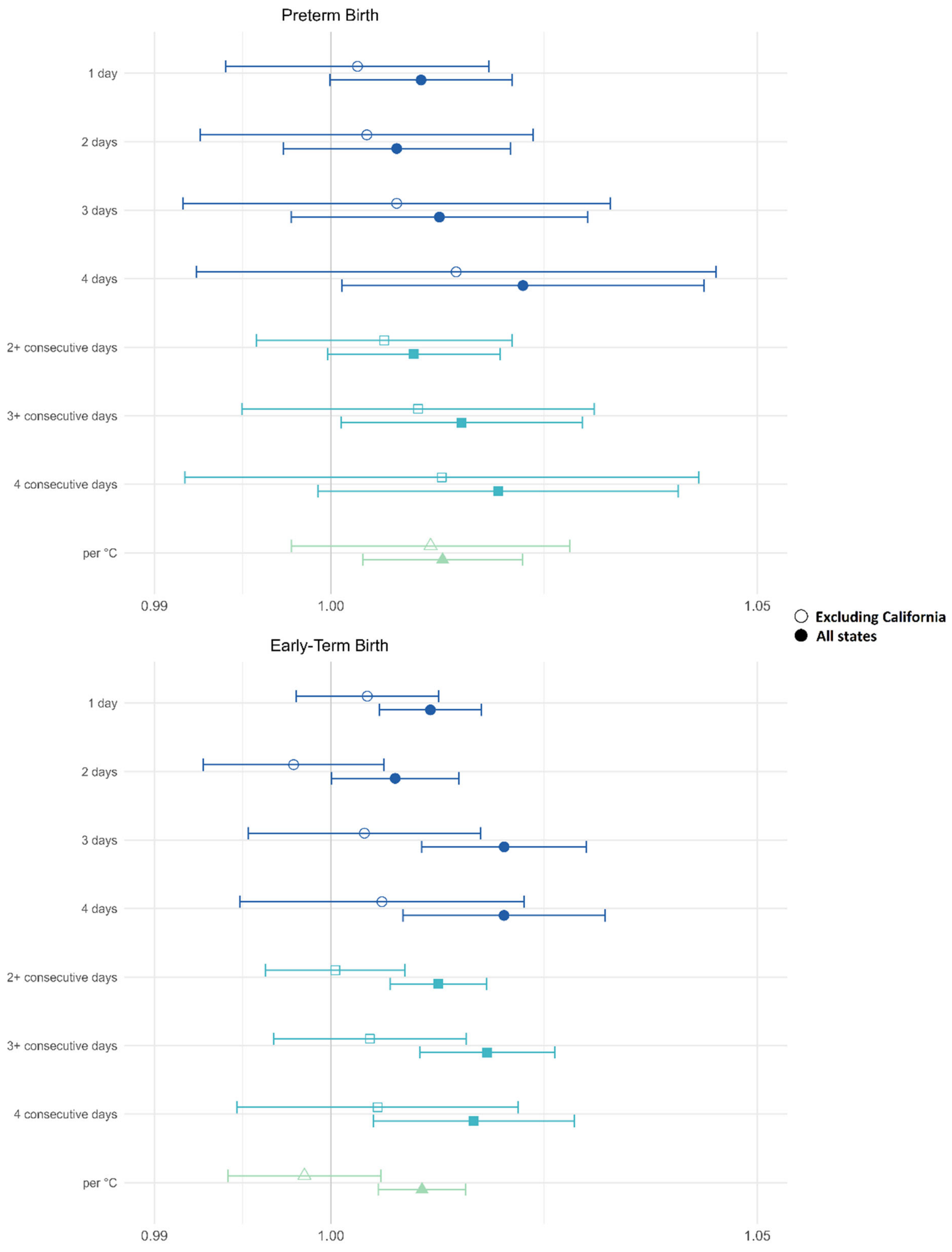


**Figure S1:** Pooled odds ratios and 95% confidence intervals for the association between heatwaves and early-term birth, including and excluding California. The reference category for heatwave definition 1 (dark blue circles) is 0 hot days in the previous week. Heatwave definition 2 (blue squares) are dichotomous exposure categories where the reference categories are <2-, <3-, and <4-consecutive days, respectively. Heatwave definition 3 (green triangle) represents the odds ratio associated with a 1°C increase in the 4-day average degrees over the 97.5th percentile.

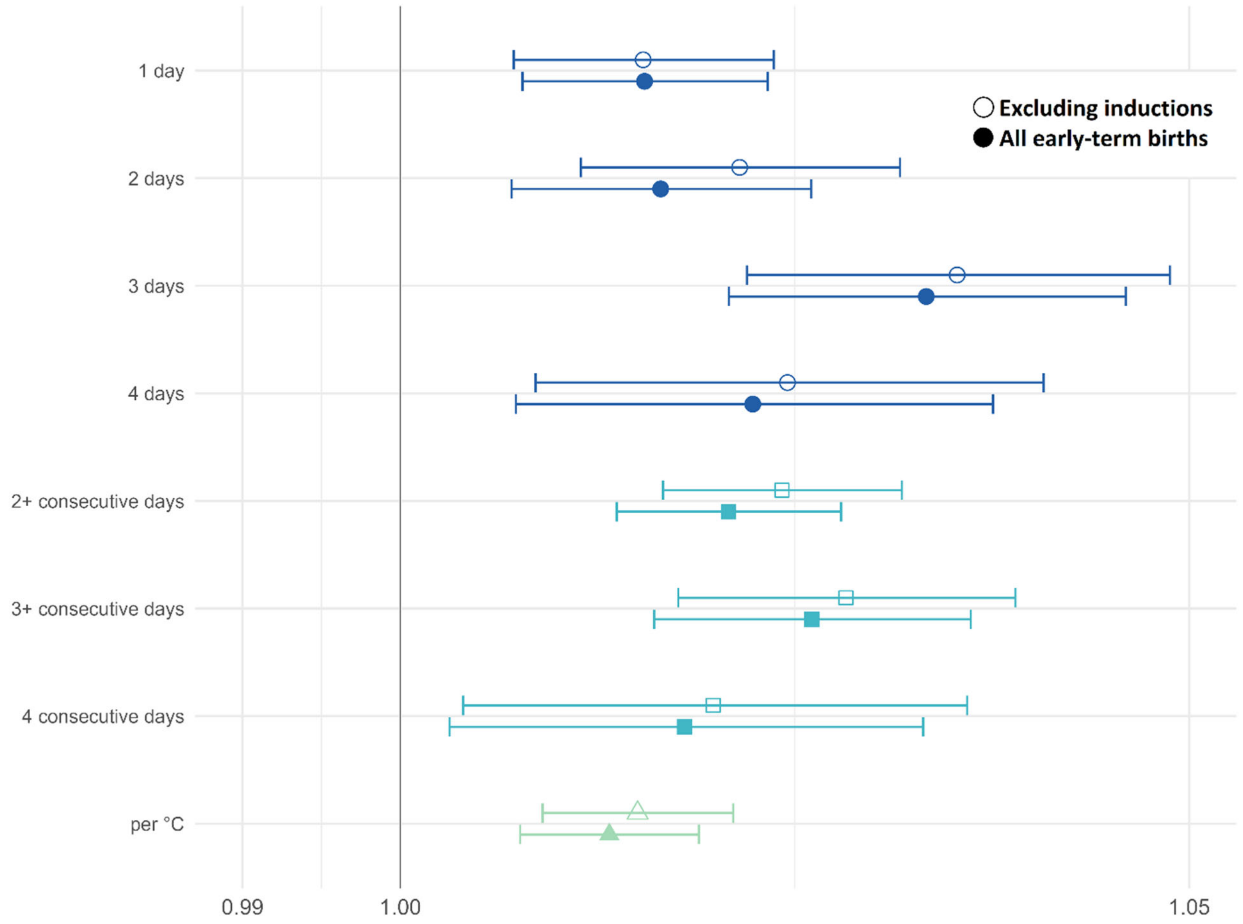


**Figure S2:** Pooled odds ratios and 95% confidence intervals for the association between heatwaves and early-term birth, including and excluding medically-induced births in California, Kansas, Nevada, and Oregon. The reference category for heatwave definition 1 (dark blue circles) is 0 hot days in the previous week. Heatwave definition 2 (blue squares) are dichotomous exposure categories where the reference categories are <2-, <3-, and <4-consecutive days, respectively. Heatwave definition 3 (green triangle) represents the odds ratio associated with a 1°C increase in the 4-day average degrees over the 97.5th percentile.

Figure S3

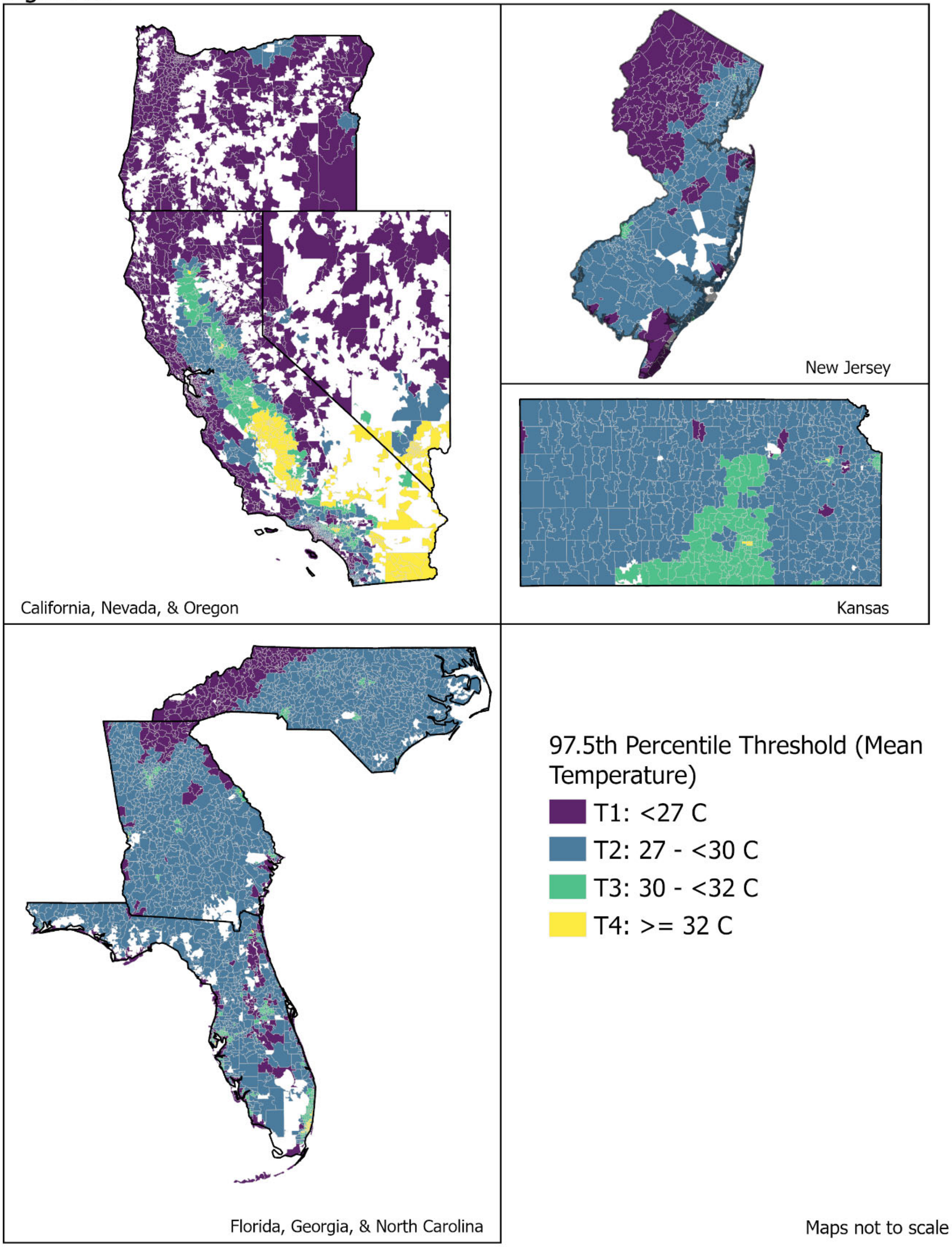


**Figure S3:** Geographic distribution of 97.5th percentile temperature categories (T1-T4) by ZCTA
